# Associations between ambient air pollutants exposure and case fatality rate of COVID-19: a multi-city ecological study in China

**DOI:** 10.1101/2020.05.06.20088682

**Authors:** Tiantian Zhang, Guoli Zhao, Li Luo, Yongzhen Li, Wenming Shi

## Abstract

**Background:** Environmental factors, including air pollution, can strongly impact on spatio-temporal patterns of infectious diseases outbreak. In this study, we aimed to investigate the association and correlation between ambient air pollutants and case fatality rate (CFR) of the novel coronavirus disease (COVID-19) in China.

**Methods:** Publicly accessible data on COVID-19 average CFR were utilized in the data analysis. The ambient daily air pollutants including fine particulate matter (PM_2.5_), inhalable particles (PM_10_) and nitrogen dioxide (NO_2_) during the period from December 25, 2019 to March 5, 2020 were obtained from National Air Quality Real-time Publishing System of China. Ecological analysis was performed to explore the association and correlation between the cumulative average exposure of ambient air pollutants at different lag days (14 and 28 days) and average CFR in China outside Hubei and cities in Hubei province via model fitting.

**Results:** The average case fatality rate was highest in Wuhan city (4.53%) and the cumulative average exposure of ambient PM_2.5_, PM_10_ and NO_2_ at lag 28 days was 55.8±12.1μg/m^3^, 66.8±9.2μg/m^3^, 20.7±4.4μg/m^3^, respectively in Hubei province during the study period. Ecological analysis showed that ambient PM_2.5_, PM_10_ and NO_2_ exposure at both lag 14 and 28 days was positively correlated with average CFR in China outside Hubei (province-level). For city-level analysis in Hubei, significant associations were only found between cumulative ambient NO_2_ exposure and average CFR(r=0.693 for Lag0-14, r=0.697 for Lag0-28, respectively) during the same period.

**Conclusion:** Our findings suggested ambient PM_2.5_, PM_10_ and NO_2_ exposure, especially at 28 lag days, positively associated with the case fatality rate of COVID-19 in China. These results could help provide guidance for identifying potential exposure window and preventing and controlling the epidemic.

## Introduction

The novel coronavirus disease (COVID-19) is an ongoing global epidemic event, which caused by severe acute respiratory syndrome coronavirus 2 (SARS-CoV-2)[1]. Respiratory droplets and human-to-human contact are the major routes of transmission of COVID-19. Some studies showed that environmental factors might influence the transmissions and risk of COVID-19[2, 3]. Since COVID-19 is a respiratory infectious disease and SARS-CoV-2 could remain viable in aerosols for hours [4], it’s interesting to investigate the associations between air pollution and COVID-19 infection, and identify possible indicator of air pollutants for blocking the epidemic. However, the study on the effects of air pollutants on the death risk of COVID-19 infection was still limited and inconsistent in China. In this study, we aimed to investigate the associations between ambient air pollutants exposure and case fatality rate (CFR) of COVID-19 in Mainland China, to provide scientific guidance for identifying potential exposure time window and preventing and controlling the epidemic.

## Methods

For this multi-city ecological study, we collected the daily cumulative confirmed and death numbers of COVID-19 cases information in China during the study period from January 22 to March 5, 2020 from the National Health Commissions (http://www.nhc.gov.cn/xcs/xxgzbd/gzbd_index.shtml) and provincial Health Commissions of China (http://wjw.hubei.gov.cn/bmdt/ztzl/fkxxgzbdgrfyyq/index.shtml). Due on no death case was reported in 5 provinces (Xizang, Qinghai, Shanxi, Jiangsu and Ningxia) by March 5, 2020 and island-province factor (Hainan), a total of 25 provinces covering 256 cities (243 cities outside Hubei, 13 main cities in Hubei) were included in our study. The average case fatality rate of COVID-19 was calculated in China outside Hubei at province-level, and in each city of Hubei province during the same period. Due on the data were all from public websites, the Ethical approval was not needed in the study.

The data on daily average concentrations of air pollutants including fine particulate matter (PM_2.5_), inhalable particles (PM_10_) and nitrogen dioxide (NO_2_) during the period from December 25, 2019 to March 5, 2020 was obtained from National Air Quality Real-time Publishing System of China. This platform provided the real-time criteria air pollutants in all state-controlled monitoring stations. To control for the potential confounders, the ambient daily average temperature, relative humidity for each city was obtained from China Meteorological Data Service Center (http://data.cma.cn/en).

The temporal cumulative average exposure of ambient PM_2_._5_, PM_10_ and NO_2_ at different lag days (14 days and 28 days) before the daily case reports was evaluated among the cities in Hubei province (city-level) and in China outside Hubei (province-level).

Ecological analysis was conducted to explore the temporal associations between the cumulative average exposure of ambient air pollutants and COVID-19 case fatality rates in China outside Hubei (province-level) and in Hubei province (city-level). Multiple regression models were fitted to the data among 24 provinces of China and city-level analysis in Hubei province alone during the study period.

## Results

Among the cities in Hubei province, the cumulative average exposure level of the ambient PM_2.5_, PM_10_ and NO_2_ at lag 28 days was 55.8±12.1μg/m^3^, 66.8±9.2 μg/m^3^, 20.7±4.4μg/m^3^, respectively during the study period. The average case fatality rate was highest in Wuhan city (4.53%), followed by Jingmen city and Ezhou city in Hubei province during the study period. For provinces outside Hubei, the average CFR was highest in Heilongjiang province and lowest in Zhejiang province of China.

The linear regression analysis showed a positive association between air pollutants exposure and average case fatality rate in China. As indicted in Figure 2, the associations are all significant between average CFR and cumulative average exposure to ambient PM_2.5_, PM_10_ and NO_2_ in both lag 14 and 28 days at province-level in China outside Hubei. The correlation coefficient of NO_2_ was highest (r=0.739, P<0.001) among the three air pollutants at lag 28 days (Figure 2). For Hubei province, city-level analysis showed a positive association between cumulative average exposure of NO_2_ and average CFR (r=0.693 for Lag0-14, r=0.697 for Lag0-28, respectively), while no significant associations were observed between ambient PM_2.5_ and PM_10_ exposure and average CFR in cities of Hubei province.

**Figure 1.**
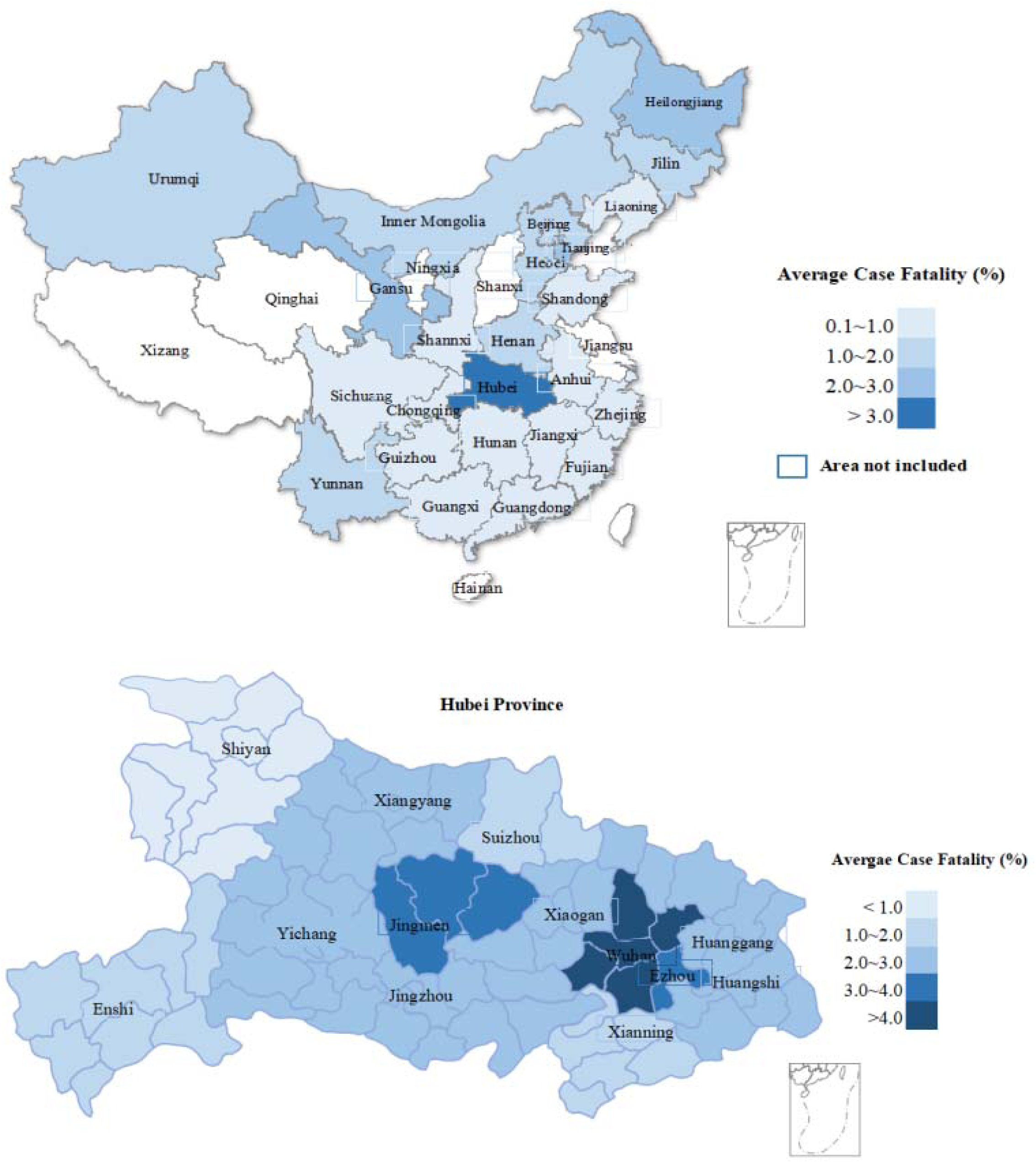
Map of the distribution of the average case fatality rate of COVID-19 in Hubei Province and in Mainland China.

**Figure 2.**
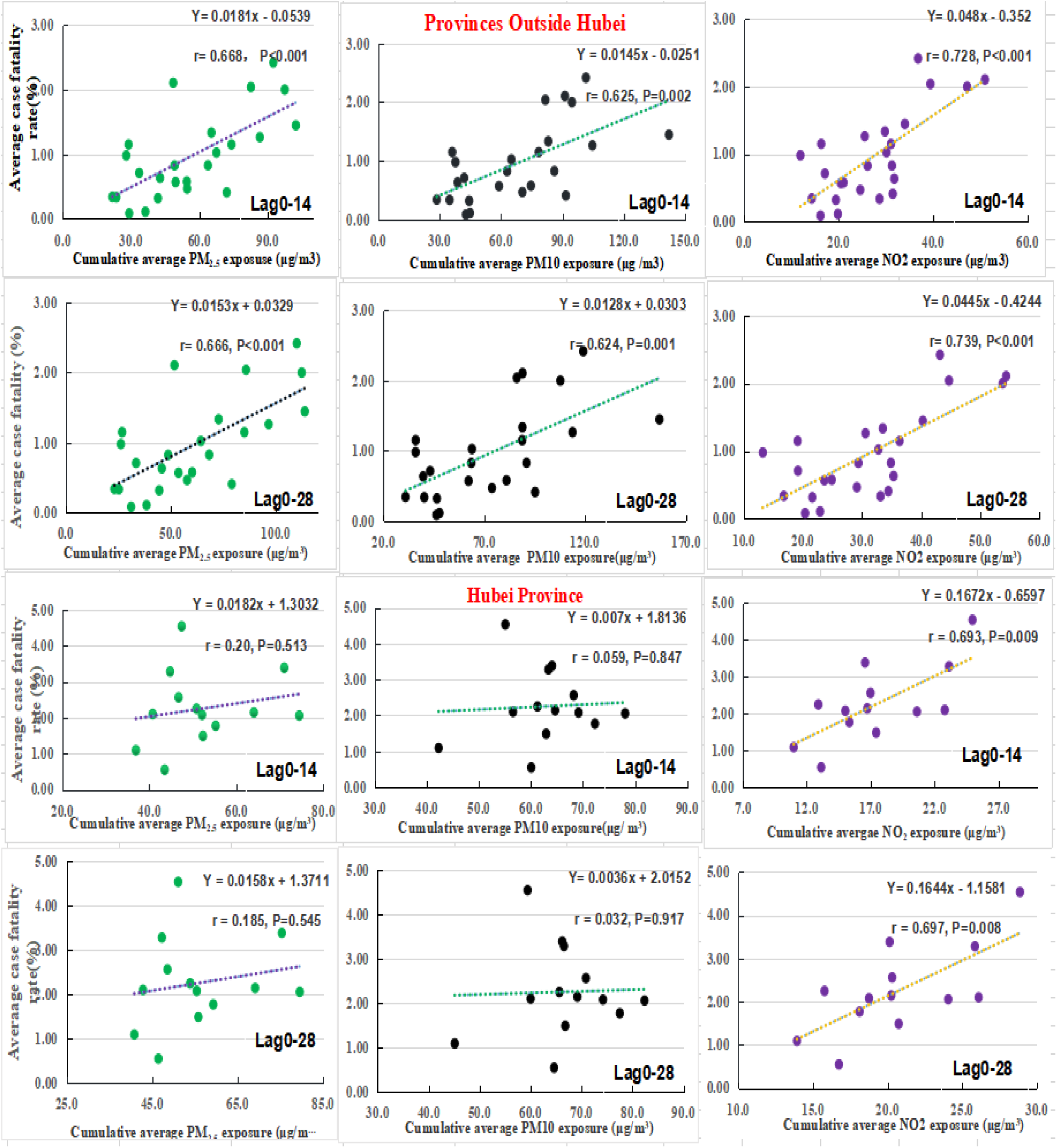
The correlation and associations between air pollutants exposure at different lag days and average case fatality rate of COVID-19 in Hubei province (city-level) and in Mainland China (province-level).

## Discussion

Our study suggested that ambient particulate matter and NO_2_ exposure was associated with the increased case fatality of COVID-19 in Mainland China. These findings could provide evidence that air pollution is an important factor in COVID-19 infections.

Previous studies has indicated that air pollution associated with a variety of respiratory disease including SARS, influenza, asthma and COPD[5–8]. A recent time-series study showed that ambient particulate matter, NO_2_ exposure was associated with the increased confirmed case number of COVID-19[9]. A study conducted in multiply countries indicated that long-term exposure to ambient NO_2_ could be a contributor factor to COVID-19 fatality[10]. Consistent with prior studies. our results indicate ambient NO_2_ exposure, especially at 28 lag days, might increase the CFR of COVID-19. The biological explanation might be that air pollutants could compromise lung function and increase COVID-19 fatality[8].

To our knowledge, this was one of the few studies performed in Chinese cities to explore the temporal associations between ambient air pollutants exposure and case fatality rate of COVID-19. The high correlation between ambient NO_2_ and CFR of COVID-19, especially at 28 lag days exposure, might help identifying which pollutants in certain exposure time window play the dominate the role in increasing the death risk of COVID-19 in China.

However, our study has some limitations. First, ecological study designs were adopted in the study, other city-level factors including implementation ability of control policy of COVID-19, the availability of medical resources might confound our findings. Second, the study period may not represent a whole air pollutants pattern associated with the average case fatality rate of COVID-19. Furthermore, the exposure misclassification might bias the study result due to the different activity patterns of people. For all these limitations, the findings should be interpreted with caution before extension to other population and research.

## Conclusions

Our study suggest positive associations between ambient PM_2.5_, PM_10_ and NO_2_ exposure and case fatality rate of COVID-19 in China. The 28-day lag effects of air pollutants can help to identify the potential exposure window that have the high adverse impact on the COVID-19 and improve the public practice for preventing the epidemic.

## Data Availability

The data were all from public websites including he National Health Commissions of China.

## Author Contributions

TZ, GZ, and WS designed the project, processed and analyzed the data, and wrote the manuscript. LL and YL collected the data and edited the manuscript. All authors critically reviewed and approved the final version of the manuscript

## Conflict of interests

The authors declare no competing financial interest.

## Acknowledgment

This study was supported by the National Natural Science Foundation of China (Grantnu:71874033).

## Notes

### Competing Interest Statement

The authors have declared no competing interest.

### Funding Statement

This study was supported by the National Natural Science Foundation of China (Grant nu: 71874033)

## References

1. Bi Q, Wu Y, Mei S, Ye C, Zou X, Zhang Z, Liu X, Wei L, Truelove SA, Zhang T, Gao W, Cheng C, Tang X, Wu X, Wu Y, Sun B, Huang S, Sun Y, Zhang J, Ma T, Lessler J, Feng T. Epidemiology and transmission of COVID-19 in 391 cases and 1286 of their close contacts in Shenzhen, China: a retrospective cohort study. LANCET INFECT DIS 2020.

2. Sobral M, Duarte GB, Da PSA, Marinho M, de Souza MA. Association between climate variables and global transmission oF SARS-CoV-2. SCI TOTAL ENVIRON 2020: 729: 138997.

3. Shi P, Dong Y, Yan H, Zhao C, Li X, Liu W, He M, Tang S, Xi S. Impact of temperature on the dynamics of the COVID-19 outbreak in China. SCI TOTAL ENVIRON 2020: 728: 138890.

4. van Doremalen N, Bushmaker T, Morris DH, Holbrook MG, Gamble A, Williamson BN, Tamin A, Harcourt JL, Thornburg NJ, Gerber SI, Lloyd-Smith JO, de Wit E, Munster VJ. Aerosol and Surface Stability of SARS-CoV-2 as Compared with SARS-CoV-1. N Engl J Med 2020: 382(16): 1564–1567.

5. Belgrave D, Granell R, Turner SW, Curtin JA, Buchan IE, Le Souef PN, Simpson A, Henderson AJ, Custovic A. Lung function trajectories from pre-school age to adulthood and their associations with early life factors: a retrospective analysis of three population-based birth cohort studies. Lancet Respir Med 2018: 6(7): 526–534.

6. Lei H, Li Y, Xiao S, Lin CH, Norris SL, Wei D, Hu Z, Ji S. Routes of transmission of influenza A H1N1, SARS CoV, and norovirus in air cabin: Comparative analyses. INDOOR AIR 2018: 28(3): 394–403.

7. Kan HD, Chen BH, Fu CW, Yu SZ, Mu LN. Relationship between ambient air pollution and daily mortality of SARS in Beijing. BIOMED ENVIRON SCI 2005: 18(1): 1–4.

8. Cui Y, Zhang ZF, Froines J, Zhao J, Wang H, Yu SZ, Detels R. Air pollution and case fatality of SARS in the People’s Republic of China: an ecologic study. Environ Health 2003: 2(1): 15.

9. Zhu Y, Xie J, Huang F, Cao L. Association between short-term exposure to air pollution and COVID-19 infection: Evidence from China. SCI TOTAL ENVIRON 2020: 727: 138704.

10. Ogen Y. Assessing nitrogen dioxide (NO2) levels as a contributing factor to coronavirus (COVID-19) fatality. SCI TOTAL ENVIRON 2020: 726: 138605.

